# Concordance of whole-genome amplified embryonic DNA with the subsequently born child

**DOI:** 10.1101/2024.01.12.24301086

**Authors:** Shenglai Li, Thomas Giardina, Maria Katz, Dhruva Chandramohan, Nathan Slotnick, Barry Behr, Noor Siddiqui, Yuntao Xia, Benjamin Podgursky

## Abstract

Before implantation subsequent to in vitro fertilization (IVF), the current options for Preimplantation Genetic Testing (PGT) are PGT for Aneuploidy (PGT-A) and, if clinically indicated, PGT for monogenic conditions (PGT-M). A more comprehensive approach involves PGT whole genome sequencing (PGT-WGS). PGT-WGS incorporates PGT-A, screens for hundreds of monogenic conditions, and can evaluate polygenic risk. Here we compare PGT-WGS results against the genome of the subsequently born child. We demonstrated high levels of concordance (both in sensitivity and precision) in exome variant calls between amplified embryonic DNA and sequenced fetal cord blood. This concordance was higher when filtering against 1300 targeted monogenic conditions implicated in birth defects, neurodevelopmental disorders, and hereditary cancer. To evaluate PGT-WGS’s ability to identify de novo variants we compared the child’s genome to parental genomes and demonstrated that PGT-WGS successfully identified 5/5 confirmed de-novo variants. We further demonstrated concordance in polygenic risk scores calculated for both the embryo and the subsequently born child. This agreement extended to both traditional polygenic scores and oligogenic scores (Type 1 diabetes, Celiac disease, and Alzheimer’s Disease), which heavily rely on accurate genotyping of HLA and APOE sites. To our knowledge, this is the first direct concordance study between a whole-genome sequencing of a trophectoderm biopsy and the DNA of the subsequently born child. By demonstrating a high degree of whole-exome concordance and adept detection of de novo variants, this approach showcases PGT-WGS’s capability to identify genetic variants not explicitly targeted for monogenic screening.

## Introduction

During an IVF cycle, embryos are commonly tested via preimplantation aneuploidy testing (PGT-A) for chromosomal abnormalities prior to implantation. This testing is performed using either a microarray designed for a restricted set of sites or low-depth next-generation sequencing. Typically, these methods identify gross chromosomal insertions or deletions exceeding 5 million base pairs^[1]^. Preimplantation aneuploidy testing for monogenic condition (PGT-M) can only be performed if the egg or sperm contributor is affected by a known genetic condition or carrier screening identifies that a couple’s children are at risk of inheriting an X-linked condition or a shared autosomal condition. Precise knowledge of the molecular specifics of the familial variant is required. In these cases, PGT-M may be performed to avoid implantation of affected embryos^[2]^.

PGT-A and PGT-M are essential for general and targeted screening of known genetic conditions. However, their goal is not to provide comprehensive genetic screening, and this leaves a significant gap in their coverage. First, few parents have undergone comprehensive genetic screening; carrier screening screens for a targeted set of autosomal recessive and X-linked conditions; many genetic conditions, most notably autosomal dominant variants, are not evaluated^[3]^. Likewise, donated embryos may have limited parental screening and personal and family history may be incomplete for the donors of sperm or eggs. De-novo variants, genetic mutations present in a child but not the father or mother, can not be identified through parental testing^[4]^. Embryos previously frozen for fertility preservation cannot be re-evaluated in light of new research without being re-biopsied, which negatively impacts implantation success rates^[5]^.

Previous research has attempted to evaluate the efficacy of whole-genome preimplantation genetic screening by comparing against born siblings and parents, attempting to statistically reconstruct the true genome of the child by identifying shared chromosomal segments^[6][7]^. This screening has until recently only been available on a research but not clinical basis^[8]^. Imputing an embryo genome has many limitations, including that it cannot detect de-novo variants in an embryo. The number of germline *de novo* variants unique to an individual is an area of active research. Commonly cited estimates suggest 1-2 exome variants among 50-100 total genomic variants per individual^[9][10]^. The contribution of these *de novo* variants to genetic disease is likewise a subject of research; current research suggests for developmental disorders alone, up to 0.5% of live births are affected by a condition caused by *de novo* mutations unique to a child^[11]^.

Since de novo variants have a substantial impact on disease pathogenicity, employing a technology to identify and characterize these variants before an affected pregnancy begins and before childbirth is highly meaningful. Orchid Health recently released the first clinically available whole-genome preimplantation screening (PGT-WGS) utilizing custom whole-genome amplification protocols^[8]^. While the precision and sensitivity of whole-genome sequencing on amplified DNA have been evaluated against known inherited variants, the ability to validate and quantify the detection of inherited variants without probe development as well as *de novo* variants has been limited until a child screened via PGT-WGS was born^[7]^. In this study, we compare and present a whole-genome sequenced embryo against the genome of the subsequently born child.

## Subjects and Methods

### Consent

Preimplantation testing and consultation was performed under IRB 20222645. Explicit consent was gathered from both parents for this study. To protect the privacy of the born child in the case of reidentification, no clinical interpretation of the embryo or child’s sequencing data is included in this study. Polygenic Risk Scores (PRS) have been masked and no exonic or variants of clinical significance are reported.

### Case

A male / female couple underwent an IVF cycle due to advancing age with an intent to perform preimplantation genetic screening. At days 6 and 7, 3 embryos (Embryo 1, Embryo 2, and Embryo 3) were deemed candidates for implantation and ∼5 cells were biopsied from the trophectoderm of each embryo for PGT. Whole-genome preimplantation genetic testing was performed by Orchid Biosciences using whole-genome amplification and sequencing. No specific monogenic testing was indicated or performed.

Three embryo biopsies were analyzed. After PGT-A analysis, two biopsies (Embryo 1, Embryo 2) were determined to be euploid and sent for higher-depth sequencing. E03 had multiple aneuploidies and further analysis was not performed.

On receipt of higher-depth sequencing data, polygenic risk scores and microduplication/deletion screening were performed on embryos Embryo 1 and Embryo 2. Both Embryo 1 and Embryo 2 were determined to be chromosomally normal. Embryo 1 was successfully implanted and a healthy child was born without complications. Prior to the birth, the parents consented and arranged for a concordance analysis between themselves, the born child, and the whole-genome biopsy to evaluate the sensitivity and precision of PGT-WGS.

### Sample collection

All embryos were created using Intracytoplasmic sperm injection (ICSI). Embryos from days 6 and 7 were biopsied following standard clinical PGT trophectoderm biopsy SOPs. Each biopsy contained approximately 5 cells and was collected in a 200ul PCR tube with 3ul of cell buffer.

Cord blood from the born child was collected immediately after delivery in EDTA tubes, and shipped on ice packs overnight. Psomagen (certified CAP #8742212, CLIA #21D2062464) performed DNA extraction.

Parental saliva samples were collected using AccuGene AccuSaliva Collection Kits and sent to the Orchid laboratory (certified CAP #9234146, CLIA #34D2260214) for extraction and preparation.

### Next-generation sequencing

Saliva and embryo samples were processed in the Orchid laboratory. Biopsies were amplified using a lab-developed protocol. DNA sizes after WGA were first confirmed by running 1-2% Agarose E-Gel (Invitrogen). Saliva gDNA was directly extracted in preparation for NGS. After size determination, 250-500ng of DNA was used for library preparation with KAPA HyperPlus kit per manufacturer’s instructions. Dual Index UMI adapters (Integrated DNA Technologies) were used in the ligation. Library concentration was quantified using the Qubit 4 dsDNA HS. Library sizes were measured through Agilent 4150 Tapestation Genomic ScreenTape assay (Agilent Technologies). Sequencing runs were performed on a MiniSeq for low-pass aneuploidy screening and by Psomagen on a NovaSeq X for 30X WGS per manufacturer’s instructions.

Cord blood was sequenced using Psomagen’s CAP/CLIA blood extraction and sequencing workflow to a depth of 30x.

### PGT-A

Preliminary aneuploidy screening was performed via Copy Number analysis on low-depth sequencing obtained from the MiniSeq. Embryos with clear (>20 million base pair) insertions or deletions were not sent for 30x sequencing. Aneuploidy was reassessed on high-depth sequencing data via NxClinical with additional screening for clinically significant microdeletions and microduplications^[12]^.

### Variant calling

Individual sample BAMs, SNP and indel variant calls were generated using the Gencove Deep-seq pipeline (Human WGS GRCH37 v1.0). Individual calling was performed to evaluate pessimistic PGT-WGS performance in the absence of gamete sources or other embryos. Joint calling was performed using a GATK best practice workflow. Joint calling was performed on all cohort embryo biopsies along with both parental samples to evaluate the performance attainable when sequencing data is available from parents and/or other embryos in an IVF retrieval cohort (typical IVF cycles produce 1-10 mature day-6 embryos)^[16]^. Variant calling was restricted to Genome-in-a-Bottle regions downloaded from https://www.nist.gov/programs-projects/genome-bottle and excludes difficult-to-map regions. Variants were filtered using best-practice internal guidelines.

### De-novo variant identification

Candidate de-novo variants were sourced from the born child. To minimize the proportion of false positive de-novo candidates in the born child, candidate variants were filtered to heterozygous variants with 0.4 < VAF < 0.6, excluding GiaB difficult regions, restricting DP to > 30 and removing indels of length 10 or greater. Variants were excluded if detected in either mother or father or if read depth was <= 15 on either parent (to minimize the possibility of allelic dropout in parent samples). Exonic variants were not evaluated, to avoid the incidental discovery of variants later found to have clinical significance for the born child.

### Sanger confirmation

Sanger sequencing was performed on amplified DNA for the confirmation of the absence or presence of suspected de-novo variants on the parents and born child. Sequencing was performed by Psomagen on extracted saliva gDNA for parents and extracted DNA from the cord blood sample for the born child. No amplified DNA remained available after 30x sequencing for Embryo 1; no confirmatory Sanger sequencing was performed.

PCR primers were designed by Sanger sequencing at Psomagen. To determine the specific position corresponding to the nucleotide of interest, BLAST was performed on each Sanger sequence to confirm unique genomic coordinates^[14]^. The genotypes of the sites of interest were determined by manual inspection of the chromatograms.

### Polygenic risk scores

Polygenic Risk Scores capturing statistical genetic risk for common diseases were built by combining publicly available summary statistics and using PRS-CS (Polygenic Prediction via Bayesian Regression and Continuous Shrinkage Priors) to obtain final disease models. PLINK 2 was used to generate scores for each sample and each disease. For some diseases, the average scores differ by ethnicity, as captured by principal components. As this does not usually reflect an actual difference in disease susceptibility, an adjusted score is created which does not differ by ethnicity. For this adjustment, the scores which are expected based on the first ten principal components for each sample were subtracted from the raw scores. Models were evaluated for Atrial Fibrillation, Alzheimer’s disease, Bipolar disorder, Coronary artery disease, Celiac disease, Inflammatory bowel disease, Schizophrenia, Type 1 diabetes, and Type 2 diabetes. The methods for developing each of these PRS are available as whitepapers at guides.orchidhealth.com.

## Results

In this study we performed a concordance study to validate three endpoints:

### Whole-genome sequencing concordance

Whole-genome variant calling on amplified embryonic DNA shows high sensitivity and precision as compared against genomic DNA from the born child.

### Detection of de-novo variants

Variant calling on amplified embryonic DNA detects de-novo variants (as identified in the born child via a parental trio analysis).

The frequency of pathogenic de-novo mutations affecting a given individual is low; on average, each child averages 1-2 de-novo exonic mutations. We do not attempt to identify pathogenic exomic de-novo variants in this born child; we instead evaluate the ability of PGT-WGS to detect de-novo mutations in non-coding regions, as a proxy for the detection of pathogenic variants during whole-genome preimplantation screening.

### Polygenic risk score stability

Polygenic predisposition screening (PGT-P) allows parents to prioritize embryos for implantation-based scores combining small effect sizes across hundreds or thousands of gene variations to produce Polygenic Risk Scores (PRS). Generally, to compute these scores, providers rely on direct testing of only a few hundred thousand sites via arrays and rely on statistical genetic imputation to reconstruct the remaining genome for testing. However, via PGT-WGS we can compute polygenic risk scores with direct measurement of the vast majority of target sites. Here we demonstrate that PRS as computed on embryo biopsies using PGT-WGS are consistent with those for the born child.

### WGS Concordance

We compared the whole-genome variant calls from the amplified DNA from Embryo 1 against the genomic DNA from the subsequently born child (hereafter, “Cord Blood”).

As this concordance was designed to measure the accuracy of PGT-WGS in a clinical context., performance was evaluated both on the overall exome and on a list of 1300 genes (Appendix A) linked to neurodevelopmental disorders, birth defects, and hereditary cancer, and as such curated for their utility during monogenic embryo screening.

Best-practice NGS QC was performed on Embryo 1 and Embryo 2 amplified DNA, parental saliva, and gDNA from the born child^[15]^. Metrics evaluated included:

- Mean cover on the exome and genome, against a target depth of 30
- Fraction of sites with a read depth of 10 or greater, on the genome, exome, and 1300 gene panel described previously, against a VAF calling threshold of depth 8.
- Fraction of aligned reads, as a measure of contamination
- Fraction of Q30 reads, indicating high-quality sequencing
- Mitochondrial content, as a measure of sample degradation

Results are shown in Table 1. Q30, GC content, and read alignment and mean cover indicated high-quality amplification and sequencing for all samples. Mitochondrial content for Embryo 2 was within expectations (healthy amplified trophectoderm biopsies typically have mitochondrial content of 2% or lower). Mitochondrial content in Embryo 1, corresponding to the born child, was higher than expected at 10%, indicating lower quality of biopsy. Despite this, 10x depth coverage still reached 96% on the 1300 gene panel identified as clinically significant and monogenic analysis was performed.

**Table 1.**
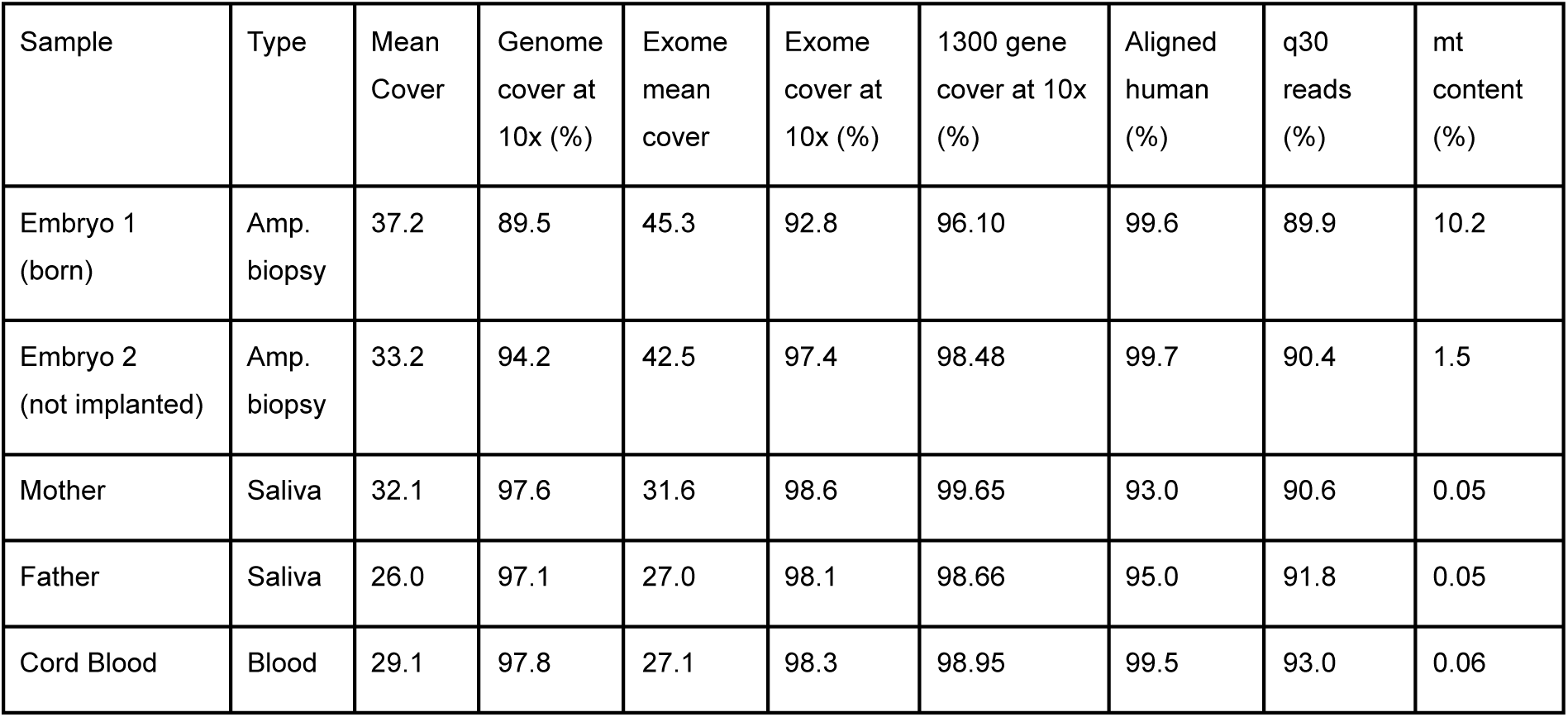
Sequencing QC Metrics. Best-practice NGS QC matrix for the amplified embryo, born child, and parental reference samples

| Sample | Type | Mean Cover | Genome cover at 10x (%) | Exome mean cover | Exome cover at 10x (%) | 1300 gene cover at 10x (%) | Aligned human (%) | q30 reads (%) | mt content (%) |
| --- | --- | --- | --- | --- | --- | --- | --- | --- | --- |
| Embryo 1 (born) | Amp. biopsy | 37.2 | 89.5 | 45.3 | 92.8 | 96.10 | 99.6 | 89.9 | 10.2 |
| Embryo 2 (not implanted) | Amp. biopsy | 33.2 | 94.2 | 42.5 | 97.4 | 98.48 | 99.7 | 90.4 | 1.5 |
| Mother | Saliva | 32.1 | 97.6 | 31.6 | 98.6 | 99.65 | 93.0 | 90.6 | 0.05 |
| Father | Saliva | 26.0 | 97.1 | 27.0 | 98.1 | 98.66 | 95.0 | 91.8 | 0.05 |
| Cord Blood | Blood | 29.1 | 97.8 | 27.1 | 98.3 | 98.95 | 99.5 | 93.0 | 0.06 |

PGT-WGS sequencing performance against true variants was evaluated next. To replicate the performance attainable during the clinical use of PGT-WGS, both individual and joint calling were performed as described in methods and materials. True baseline variants were identified by filtering for variants present in the cord blood and at least one parent, to eliminate false positives present in the cord blood, and filtered according to the scope of variants described in methods and materials.

Table 2 shows the resulting concordance as measured across the exome and 1300 gene screening panel for each of individual and joint calling. Calling sensitivity and precision on the 1300 gene screening panel was higher than overall exomic calls, with precision of 99.2% at 96.7% sensitivity. In all cases, joint calling improved sensitivity at the cost of precision (on the screening panel, a gain of .6% sensitivity resulted in 1.5% loss of precision).

**Table 2.** Variant calling sensitivity and precision. NGS calling performance on Embryo 1 against the subsequently born child’s cord blood.

|  | True Positives | False Positives | False Negatives | Precision | Sensitivity | F-measure |
| --- | --- | --- | --- | --- | --- | --- |
| 1300 gene (joint) | 1385 | 32 | 37 | 97.74% | 97.40% | 97.57% |
| Exome (joint) | 14095 | 425 | 526 | 97.07% | 96.40% | 96.74% |
| 1300 gene (individual) | 1376 | 11 | 46 | 99.21% | 96.77% | 97.97% |
| Exome (individual) | 13927 | 195 | 694 | 98.62% | 95.25% | 96.91% |

### De-Novo Variant Detection

We next performed a retrospective analysis to determine whether PGT-WGS at 30x read depth detected de-novo variants which were later confirmed on the born child via trio analysis against parental DNA. Using the filters described in the methods above on the born child variants resulted in 15 SNP and 1 deletion de-novo candidates. Due to limited availability of DNA, 4 SNP sites were randomly sampled along with the one deletion for confirmatory Sanger sequencing (chromatograms available as Appendix B). Sanger sequencing was also performed on parent samples to confirm ref/ref status. All 5 of the selected variants were confirmed as de-novo mutations via confirmatory Sanger sequencing on the mother, father, and born child.

The WGS calls for Embryo 1 are shown in Table 3. All 5 variants were confirmed to have been detected during preimplantation whole-genome sequencing. IGV plots for embryo, blood, and parent WGS calls available as Appendix C.

**Table 3.** Detection of de-novo variants. WGS and Sanger genotypes for de-novo variants in parents, cord blood, and embryo. WGS calls refer to direct variant identification; no imputation or karyomapping from parents or proband was performed.

| Gene | Variant (hg19) | Mother<br>WGS | Mother<br>Sanger | Father<br>WGS | Father<br>Sanger | Cord Blood<br>WGS | Cord Blood<br>Sanger | Embryo 1<br>WGS |
| --- | --- | --- | --- | --- | --- | --- | --- | --- |
| *IL31RA*<br>(intronic) | chr5:55170862 GA⇒G | 0/0 | 0/0 | 0/0 | 0/0 | 0/1 | 0/1 | **0/1** |
|  | chr4:31632725 G⇒A | 0/0 | 0/0 | 0/0 | 0/0 | 0/1 | 0/1 | **0/1** |
|  | chr4:78544146 G⇒A | 0/0 | 0/0 | 0/0 | 0/0 | 0/1 | 0/1 | **0/1** |
|  | chr12:54176997 C⇒T | 0/0 | 0/0 | 0/0 | 0/0 | 0/1 | 0/1 | **0/1** |
| *ENSG00000231121*<br>(intronic) | chr12:77921310 C⇒T | 0/0 | 0/0 | 0/0 | 0/0 | 0/1 | 0/1 | **0/1** |

### PRS Concordance

Last we evaluated whether Polygenic Risk Scores for the 10 diseases evaluated remained consistent between the embryo biopsy and the born child. The results are shown in Figure 1. The average discrepancy in population PRS percentile between the born child and embryo was 2.4%, with a maximum discrepancy of 6.4%.

**Figure 1.**
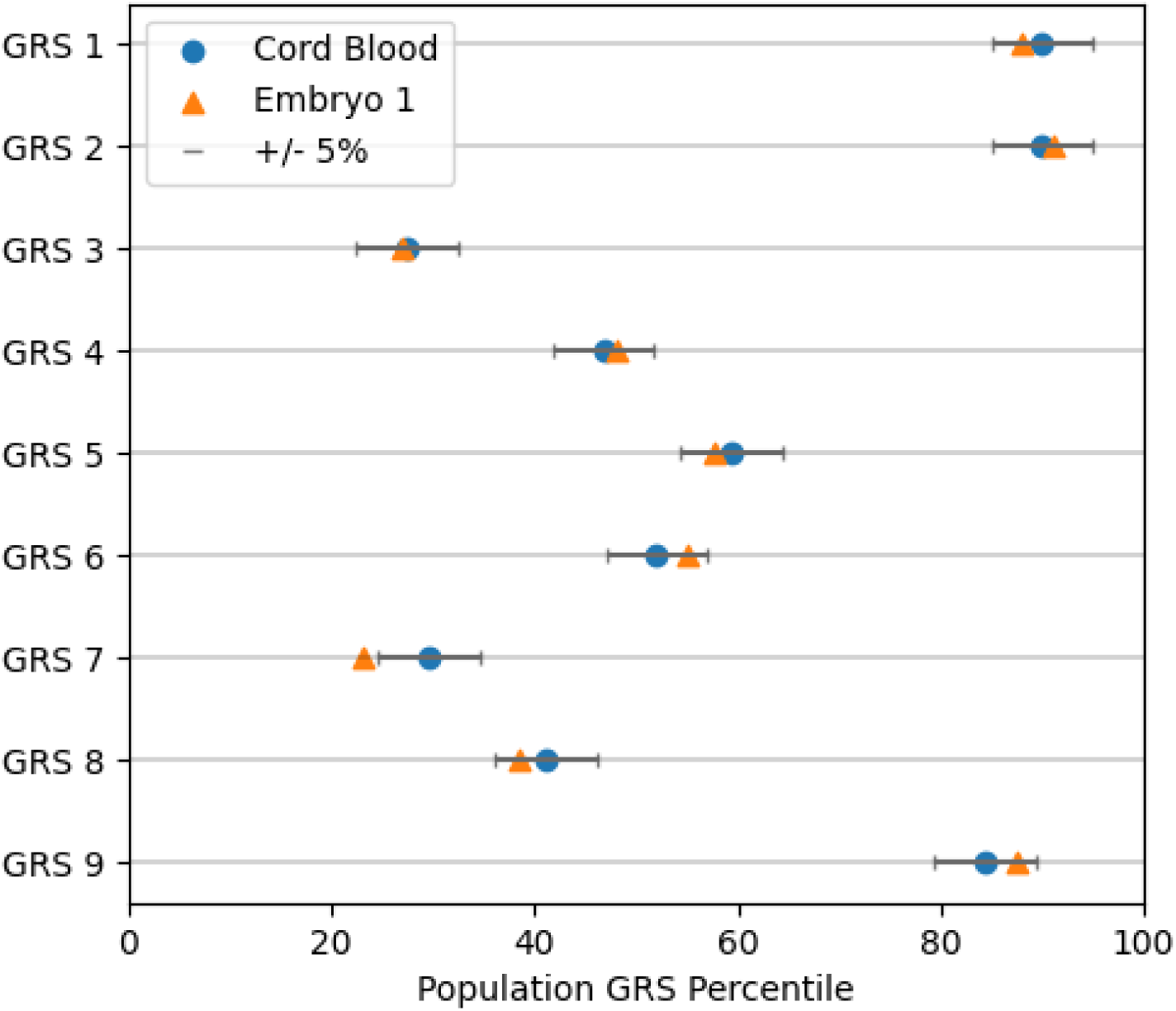
Cord Blood and Embryo GRS Concordance. Comparison of population risk percentile on 9 evaluated PRS between embryo and born child.

While most Polygenic Risk Scores are highly polygenic and do not rely on highly accurate genotyping of each individual variant site, 3 of the evaluated PRS have significant monogenic contributions and as such are deemed Oligogenic — certain HLA sites for Celiac disease, the Type 1 diabetes PRS, and the APOE sites for Alzheimer’s disease^[17]^ We validated that the WGS sequencing for Embryo 1 was consistent with the born child cord blood on all oligogenic sites evaluated in these PRS, as shown in Table 4. All Embryo 1 WGS genotypes at the relevant high-impact oligogenic sites were concordant with the genotype of the born child.

**Table 4.** HLA and APOE Site Concordance. Concordance between Embryo 1 and Cord Blood on variants relevant to 3 oligogenic PRS. Genotype is masked to preserve privacy.

| SNP RSID | Embryo 1 sequencing depth | Embryo 1 matches cord blood genotype |
| --- | --- | --- |
| APOE (ALZHEIMERS) |  |  |
| rs7412 | 64 | YES |
| rs429358 | 62 | YES |
| HLA (CELIAC) |  |  |
| rs1049124 | 54 | YES |
| rs2187668 | 24 | YES |
| rs73405471 | 48 | YES |
| rs9275437 | 17 | YES |
| HLA (TYPE 1 DIABETES) |  |  |
| rs9275437 | 17 | YES |
| rs17843689 | 42 | YES |
| rs9273369 | 88 | YES |
| rs17211699 | 83 | YES |
| rs111485156 | 41 | YES |
| rs10947332 | 28 | YES |
| rs1281935 | 62 | YES |
| rs62406889 | 24 | YES |
| rs28746898 | 28 | YES |
| rs12527228 | 19 | YES |
| rs3129727 | 12 | YES |
| rs9405117 | 37 | YES |
| rs16822632 | 43 | YES |
| rs117806464 | 93 | YES |

## Discussion

To our knowledge this is the first direct comparison of a 30x whole genome sequenced embryo against the subsequently born child. Consequently, when interpreting these results it is important to consider the constraints inherent in evaluating a single embryo/child pair; further comparisons between biopsies and born children will provide evidence regarding the consistency and replicability of these findings. It is noteworthy that the biopsy assessed in this study experienced an unusually high degree of degradation before sequencing.

Although our methodology successfully identified all de-novo variants through whole-genome sequencing, it is important to acknowledge the inherent difficulty in quantifying such variants, even in adults^[18]^. Ongoing efforts are directed towards estimating the yield of PGT-WGS in detecting de-novo variants. The filtering process, which focused on candidate de-novo variants with high confidence in the born child, may have resulted in an over-representation of variants where PGT-WGS exhibited similarly high sensitivity. A more accurate estimation of the yield of PGT-WGS on de-novo variants can be achieved through higher-depth sequencing on the born child and Sanger confirmation on a broader spectrum of candidate de-novo variants.

The concordance observed validates the high sensitivity and precision of PGT-WGS, particularly when applied to the curated list of genes chosen for screening based on their significance in birth and developmental outcomes. Overall exome precision and sensitivity are high for the evaluated variants, and Polygenic Risk Scores remain consistent between biopsy sequencing and the born child. Most significantly, PGT-WGS captured all confirmed de-novo variants, a capability not possible via any other existing form of preimplantation screening.

These results give evidence of the clinical utility of whole-genome preimplantation sequencing in detecting not only inherited monogenic conditions and genetic predispositions but also de-novo variants which would not be detected via standard preimplantation screening options.

## Data Availability

There are restrictions to the availability of the sequencing data analyzed in this study due to concerns for the privacy and confidentiality of the sequenced child and parents. Subsets of the data may be available from the corresponding authors on request.

## Acknowledgments

Funding for this research was provided by Orchid Health. We would like to thank Psomagen (CAP & CLIA) for the sequencing services.

## Author contributions

Investigation performed by SL and TG. Draft written by BP. Conceptualization of research project from BB, Noor S, Nathan S, MK, and YX. Patient consultation and sample collection by MK. Amplification methodology and lab investigation performed by YX. Review and editing by SL, YX, TG, MK, and DC.

## Appendices

### Appendix A Screened list of 1300 genes, selected due to associations with neurodevelopmental disorders, birth defects or hereditary cancer

*A2ML1,ABAT,ABCA12,ABCA3,ABCA4,ABCB11,ABCB4,ABCC2,ABCC4,ABCC8,ABCD1,ABCG5,ABCG8,ABHD12,ACAD8,ACAD9,ACADM,ACADS,ACADSB,ACADVL,ACAT1,ACOX1,ACOX2,ACSF3,ACTA1,ACTA2,ACTB,ACTC1,ACTG1,ACTN1,ACTN2,ACVRL1,ACY1,ADA,ADAMTS13,ADAMTS18,ADAMTS2,ADAMTSL4,ADNP,ADSL,AFF2,AGA,AGL,AGXT,AHCY,AHDC1,AHI1,AICDA,AIFM1,AIMP1,AIPL1,AIRE,AK2,AKR1D1,AKT3,ALAD,ALAS2,ALDH18A1,ALDH3A2,ALDH4A1,ALDH5A1,ALDH7A1,ALDOB,ALG1,ALG13,ALG6,ALG9,ALK,ALMS1,ALPK3,ALPL,AMACR,AMN,AMT,ANK1,ANK2,ANK3,ANKRD11,ANKS6,ANO10,ANO6,ANXA11,AP1S1,AP4B1,AP4E1,AP4M1,APC,APOB,APOL1,AQP2,AQP5,AR,ARG1,ARHGEF9,ARID1A,ARID1B,ARL6,ARPC1B,ARSA,ARSB,ARX,ASL,ASNS,ASPA,ASS1,ASXL1,ASXL2,ATF6,ATL1,ATL3,ATM,ATP13A2,ATP1A2,ATP1A3,ATP6AP2,ATP6V1B1,ATP7A,ATP7B,ATP8B1,ATRX,AUTS2,AVPR2,BAAT,BAG3,BAP1,BARD1,BBS1,BBS10,BBS12,BBS2,BBS4,BBS5,BBS7,BBS9,BCKDHA,BCKDHB,BCKDK,BCL11A,BCOR,BCS1L,BIN1,BLM,BLNK,BLOC1S3,BMP1,BMPER,BMPR1A,BRAF,BRCA1,BRCA2,BRIP1,BRSK2,BRWD3,BSCL2,BSND,BTD,BTK,BUB1B,C1QB,CA5A,CABP2,CACNA1C,CACNA1D,CACNA1S,CAD,CALM1,CALM2,CALM3,CANT1,CAPN3,CAPN5,CARD11,CASK,CASQ2,CASR,CAT,CBL,CBS,CC2D2A,CCDC103,CCDC40,CCDC88C,CCN6,CD247,CD3D,CD3E,CD3G,CD40,CD40LG,CD59,CD79A,CD79B,CDAN1,CDC42,CDC73,CDH1,CDH23,CDH3,CDK13,CDK4,CDK5RAP2,CDKL5,CDKN1B,CDKN2A,CEACAM16,CEBPA,CEL,CENPJ,CEP152,CEP164,CEP290,CEP57,CEP78,CERKL,CFTR,CHAT,CHD2,CHD7,CHD8,CHEK2,CHRNE,CHRNG,CIB2,CIITA,CISD2,CLCN1,CLCN4,CLCNKB,CLDN14,CLN3,CLN5,CLN6,CLN8,CLPP,CLRN1,CNGA3,CNGB3,CNKSR2,CNNM4,CNOT3,CNTNAP2,COA7,COCH,COL11A2,COL17A1,COL1A1,COL27A1,COL2A1,COL3A1,COL4A3,COL4A4,COL5A1,COL6A2,COL6A3,COL7A1,COQ4,CORO1A,COX10,COX15,COX20,COX6B1,CPS1,CPT1A,CPT2,CR2,CRADD,CRB1,CRB2,CRBN,CREBBP,CRTAP,CSNK2B,CSRP3,CSTB,CTCF,CTH,CTLA4,CTNNB1,CTNS,CTSA,CTSC,CTSF,CTSK,CUL3,CUL4B,CYBA,CYBB,CYLD,CYP11A1,CYP11B1,CYP11B2,CYP17A1,CYP19A1,CYP1B1,CYP21A2,CYP27A1,CYP27B1,CYP4V2,CYP7B1,DAG1,DBT,DCAF17,DCLRE1C,DCX,DDB2,DDC,DDR2,DDX11,DDX3X,DDX41,DEPDC5,DES,DGAT1,DGUOK,DHCR24,DHCR7,DHDDS,DHTKD1,DIAPH1,DICER1,DIS3L2,DKC1,DLAT,DLD,DLG3,DLL3,DMD,DNAH1,DNAH11,DNAH5,DNAH8,DNAI1,DNAI2,DNAJB11,DNAJB2,DNAJC12,DNAJC19,DNAJC5,DNM1,DNM1L,DNM2,DNMT1,DNMT3B,DOCK6,DOK7,DOLK,DSC2,DSG2,DSP,DST,DTNBP1,DUOX2,DUOXA2,DYNC2H1,DYRK1A,DYSF,DZIP1L,EARS2,ECHS1,EDA,EDN3,EFEMP1,EFEMP2,EFNB1,EGFR,EGR2,EHMT1,EIF2AK3,EIF2B1,EIF2B2,EIF2B3,EIF2B4,ELANE,ELP1,ENG,EOGT,EP300,EPB42,EPCAM,EPG5,EPM2A,ERBB3,ERCC2,ERCC3,ERCC4,ERCC5,ERCC6,ERCC8,ERF,ESPN,ESRRB,ETFA,ETFDH,ETV6,EVC,EVC2,EXOSC3,EXT1,EXT2,EYA1,EYA4,EYS,F10,F11,F12,F13A1,F2,F5,F8,F9,FAH,FAM161A,FANCA,FANCC,FANCD2,FANCE,FANCF,FANCG,FANCI,FANCL,FARS2,FBN1,FBP1,FBXL4,FBXO7,FERMT3,FGA,FGB,FGD1,FGD4,FGFR1,FGFR2,FGFR3,FH,FKBP10,FKRP,FKTN,FLAD1,FLCN,FLI1,FLNA,FLNC,FLVCR1,FMO3,FMR1,FOLR1,FOXC1,FOXE3,FOXG1,FOXN1,FOXP1,FOXP2,FRAS1,FREM2,FRMD4A,FTSJ1,FUCA1,FXN,G6PC1,G6PC3,G6PD,GAA,GABRA1,GABRB3,GABRG2,GAD1,GALC,GALE,GALNS,GALNT3,GALT,GAMT,GAN,GANAB,GATA1,GATA2,GATA3,GATAD2B,GATM,GBA,GBE1,GCDH,GCH1,GCK,GDAP1,GDF2,GDF5,GDI1,GFAP,GFI1B,GFM1,GFM2,GGCX,GHR,GHRHR,GJB1,GJB2,GJB3,GJB6,GLA,GLB1,GLDC,GLE1,GLS,GLUD1,GLUL,GNAI1,GNAO1,GNB4,GNE,GNPAT,GNPTAB,GNPTG,GNS,GORAB,GP1BA,GP6,GP9,GPC3,GPSM2,GPT2,GREM1,GRHPR,GRIA3,GRIK2,GRIN1,GRIN2A,GRIN2B,GRIN2D,GRIP1,GRK1,GRXCR1,GRXCR2,GSS,GSTZ1,GUCY2D,GUSB,GYS2,HAAO,HACD1,HADH,HADHA,HADHB,HAMP,HAX1,HBA1,HBA2,HBB,HCFC1,HDAC8,HEXA,HEXB,HFE,HGD,HGSNAT,HINT1,HJV,HLCS,HMGCL,HMGCS2,HMOX1,HNF1A,HNF1B,HNF4A,HNRNPU,HOGA1,HOMER2,HOXA1,HPD,HPDL,HPRT1,HPS1,HPS3,HPS4,HPS5,HPS6,HRAS,HRG,HSD17B10,HSD17B3,HSD3B2,HSD3B7,HSPB8,HSPG2,HTRA2,HUWE1,HYAL1,HYDIN,HYLS1,IDS,IDUA,IFT122,IGHM,IGHMBP2,IGLL1,IGSF1,IKBKB,IKBKG,IL1RAPL1,IL2RG,IL7R,ILDR1,INVS,IQSEC2,ITGA2B,ITGA6,ITGB3,ITGB4,ITK,ITPR1,IVD,IYD,JAK3,JPH2,KANSL1,KAT6A,KBTBD13,KCNA2,KCNB1,KCNE1,KCNH2,KCNJ1,KCNJ11,KCNK3,KCNMA1,KCNQ1,KCNQ2,KCNQ3,KCNQ4,KCNT1,KCTD7,KDM5B,KDM5C,KDM6B,KDR,KIF11,KIF1A,KIF21A,KIF5A,KIF5C,KIT,KLHL40,KLHL41,KMT2A,KMT2D,KMT5B,KNG1,L1CAM,LAMA2,LAMA3,LAMB1,LAMB3,LAMC2,LAMP2,LARGE1,LARS2,LAT,LCK,LDLR,LDLRAP1,LGI1,LHFPL5,LHX3,LIPA,LIPT1,LITAF,LMNA,LMX1B,LOXHD1,LPAR6,LPIN1,LPL,LRAT,LRP2,LRPPRC,LRTOMT,LZTR1,MAGEL2,MAK,MAN2B1,MANBA,MAOA,MAP2K1,MAPKBP1,MARVELD2,MAT1A,MATR3,MAX,MBD5,MBTPS2,MCCC1,MCCC2,MCFD2,MCOLN1,MCPH1,MECOM,MECP2,MECR,MED12,MED13L,MED17,MEF2C,MEFV,MEGF10,MEGF8,MEN1,MESP2,MET,MFAP5,MFF,MID1,MIR96,MKKS,MKS1,MLC1,MLH1,MLH3,MLYCD,MMAA,MMAB,MMACHC,MOCS1,MOCS2,MOGS,MPI,MPL,MPV17,MPZ,MRAS,MRE11,MSH2,MSH3,MSH6,MSRB3,MSX2,MT-ATP6,MT-ND1,MT-ND4,MT-ND5,MT-ND6,MT-TK,MT-TV,MTAP,MTHFR,MTM1,MTMR2,MTOR,MTR,MTRR,MTTP,MUSK,MUTYH,MVK,MYBPC3,MYH11,MYH14,MYH2,MYH7,MYH9,MYL2,MYL3,MYLK,MYO18B,MYO5A,MYO7A,MYPN,MYT1L,NAA10,NAGA,NAGLU,NAGS,NBEA,NBEAL2,NBN,NCF2,NDP,NDRG1,NDUFA1,NDUFA11,NDUFA13,NDUFA2,NDUFA9,NDUFAF5,NDUFAF6,NDUFB10,NDUFB11,NDUFB8,NDUFC2,NDUFS2,NDUFS3,NDUFS6,NDUFS8,NDUFV1,NEB,NEDD4L,NEFH,NEK8,NEU1,NF1,NF2,NFKB1,NFKB2,NHEJ1,NHLRC1,NHS,NIPBL,NLGN3,NLGN4X,NMNAT1,NOD2,NPC1,NPC2,NPHP3,NPHP4,NPHS1,NPHS2,NR0B1,NR2E3,NR3C2,NR4A2,NRAS,NRXN1,NSD1,NSD2,NSDHL,NSMCE3,NSUN2,NTRK1,NUBPL,NYX,OAT,OCA2,OCRL,OFD1,OGT,OPA1,OPA3,ORAI1,OSBPL2,OSTM1,OTC,OTOA,OTOG,P2RX2,P3H1,PACS1,PAH,PAK3,PALB2,PANK2,PAX2,PAX3,PAX7,PC,PCBD1,PCCB,PCDH15,PCDH19,PCGF2,PCNT,PCSK9,PCYT1A,PDGFRA,PDHA1,PDHB,PDSS2,PDX1,PDZD7,PEPD,PET100,PEX10,PEX11B,PEX12,PEX14,PEX16,PEX19,PEX2,PEX26,PEX5,PEX6,PFKM,PFN1,PGM3,PHF6,PHF8,PHGDH,PHKB,PHKG2,PHOX2B,PHYH,PIGA,PIGN,PIGO,PIK3CA,PIK3CD,PIK3R1,PIK3R2,PIP5K1C,PJVK,PKD2,PKHD1,PKP2,PLA2G6,PLAA,PLAU,PLEKHG5,PLG,PLOD1,PLP1,PMM2,PMS2,PNKP,PNPO,POC1A,POGZ,POLG,POLH,POLR2A,POMGNT1,POMGNT2,POMT1,POMT2,PORCN,POU1F1,POU3F4,POU4F3,PPM1D,PPP1CB,PPP2R5D,PPT1,PQBP1,PRCD,PRDM5,PRF1,PRICKLE1,PRKAG2,PRKAR1A,PRKCSH,PRKDC,PROC,PRODH,PROP1,PRPS1,PSAP,PSAT1,PSPH,PTCH1,PTCHD1,PTEN,PTPN11,PUF60,PURA,PUS1,PYCR1,PYGL,PYGM,QDPR,RAB23,RAB27A,RAB39B,RAB3GAP1,RAB7A,RAD21,RAD51C,RAF1,RAG1,RAG2,RAI1,RAPSN,RARS2,RASGRP2,RB1,RBM20,RBM8A,RCBTB1,RDH12,RDH5,RDX,RECQL4,REN,RET,RGS9,RHAG,RHBDF2,RLBP1,RMRP,RNASEH2A,RNASEH2B,RNASEH2C,ROGDI,RPE65,RPGR,RPGRIP1L,RPL10,RPS10,RPS24,RPS6KA3,RS1,RSPH1,RSPH4A,RSPH9,RTEL1,RUNX1,RYR1,RYR2,S1PR2,SACS,SAMD9,SAMHD1,SARS2,SATB2,SBDS,SBF1,SBF2,SCARB2,SCN11A,SCN1A,SCN1B,SCN2A,SCN3A,SCN4A,SCN5A,SCN8A,SCNN1A,SCNN1G,SCO1,SCO2,SDHAF2,SDHB,SDHC,SDHD,SEC23B,SEC61A1,SEC63,SEPSECS,SERAC1,SERPINA1,SERPINB6,SERPINC1,SERPIND1,SERPINE1,SERPINF2,SET,SETBP1,SETX,SGCA,SGCD,SGSH,SH3TC2,SHANK2,SHANK3,SKI,SKIV2L,SLC12A1,SLC12A3,SLC12A6,SLC16A2,SLC17A5,SLC19A2,SLC19A3,SLC1A2,SLC1A3,SLC1A4,SLC22A5,SLC25A13,SLC25A15,SLC25A20,SLC25A4,SLC25A46,SLC26A2,SLC26A3,SLC26A4,SLC27A4,SLC2A1,SLC2A2,SLC34A3,SLC35A2,SLC35A3,SLC37A4,SLC38A8,SLC39A4,SLC39A7,SLC3A1,SLC45A2,SLC4A1,SLC4A11,SLC4A3,SLC52A2,SLC52A3,SLC5A5,SLC5A7,SLC6A1,SLC6A19,SLC6A8,SLC7A7,SLC7A9,SLC9A6,SLFN14,SLITRK6,SMAD2,SMAD3,SMAD4,SMAD9,SMARCA2,SMARCA4,SMARCAL1,SMARCB1,SMARCE1,SMC1A,SMC3,SMCHD1,SMN1,SMN2,SMPD1,SMPX,SMS,SNAP29,SNRNP200,SNX10,SOD1,SOX10,SOX5,SPEG,SPG11,SPR,SPTAN1,SPTBN4,SRC,SRD5A2,SSBP1,ST3GAL3,ST3GAL5,STAC3,STAR,STAT3,STIM1,STK11,STK4,STRC,STX11,STXBP1,STXBP2,SUCLG1,SUFU,SUMF1,SUOX,SURF1,SYN1,SYNE4,SYNGAP1,SYNJ1,SYP,SZT2,TACO1,TANGO2,TAOK1,TARDBP,TAT,TBCD,TBCE,TBL1XR1,TBR1,TBX19,TBX4,TBX5,TBXAS1,TCF12,TCF20,TCF4,TCF7L2,TCIRG1,TCN2,TCOF1,TECPR2,TECTA,TERT,TF,TFR2,TG,TGFB2,TGFBR1,TGFBR2,TGM1,THBD,THPO,TIMM50,TIMM8A,TIMP3,TK2,TMC1,TMEM126B,TMEM127,TMEM216,TMEM231,TMEM237,TMEM43,TMIE,TMPRSS3,TNFRSF13B,TNNC1,TNNI3,TNNT2,TNRC6B,TNXB,TP53,TPK1,TPM1,TPM2,TPM3,TPM4,TPO,TPP1,TRAPPC9,TRDN,TREX1,TRHR,TRIM32,TRIM8,TRIO,TRMU,TRPM6,TRPV4,TSC1,TSC2,TSEN54,TSFM,TSHB,TSHR,TSPAN7,TTC19,TTC37,TTC7A,TTN,TTPA,TTR,TUBA4A,TUBB1,TUBB2B,TUBB4B,TULP1,TYMP,TYR,TYRP1,UBE2A,UBE3A,UBE3B,UBQLN2,UBR1,UNC13D,UNC80,UNG,UPF3B,UQCRC2,USH1C,USH1G,USH2A,USP7,VAPB,VCAN,VCL,VCP,VDR,VHL,VIPAS39,VKORC1,VLDLR,VPS11,VPS13A,VPS33B,VPS35,VPS45,VPS53,VRK1,VSX2,VWF,WAC,WAS,WDR45,WDR62,WFS1,WHRN,WNT10A,WRAP53,WRN,WT1,WWTR1,XPA,XPC,XPNPEP3,ZAP70,ZBTB24,ZC4H2,ZDHHC9,ZEB2,ZFYVE26,ZMPSTE24,ZNF292,ZNF462,ZNF469,ZNF711*

### Appendix B Sanger chromatograms for confirmation of de-novo variants in the born child and ref/ref genotype in parents

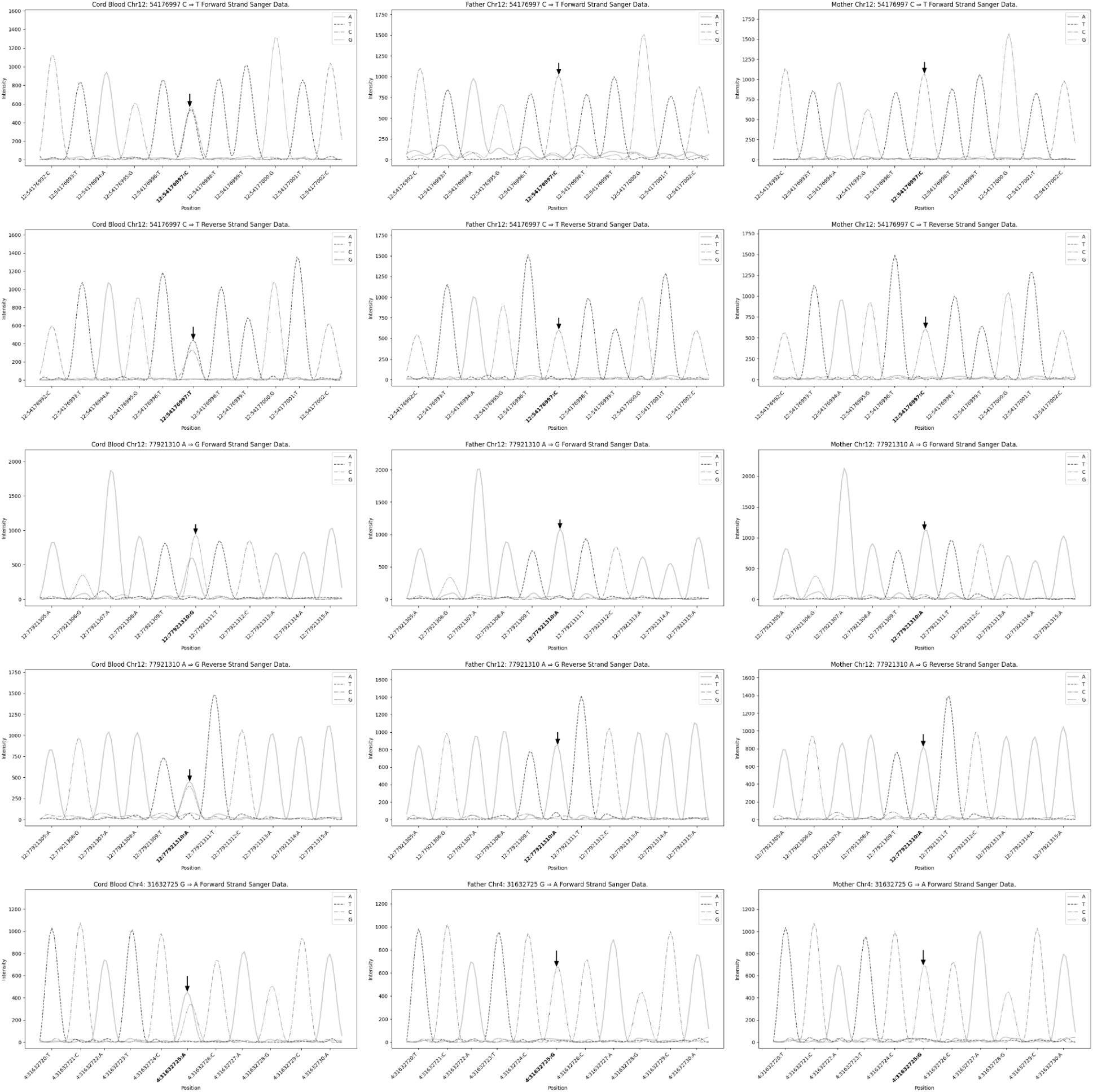

### Appendix C IGV plots for sampled de-novo variants in embryo 1, mother, father, and cord blood

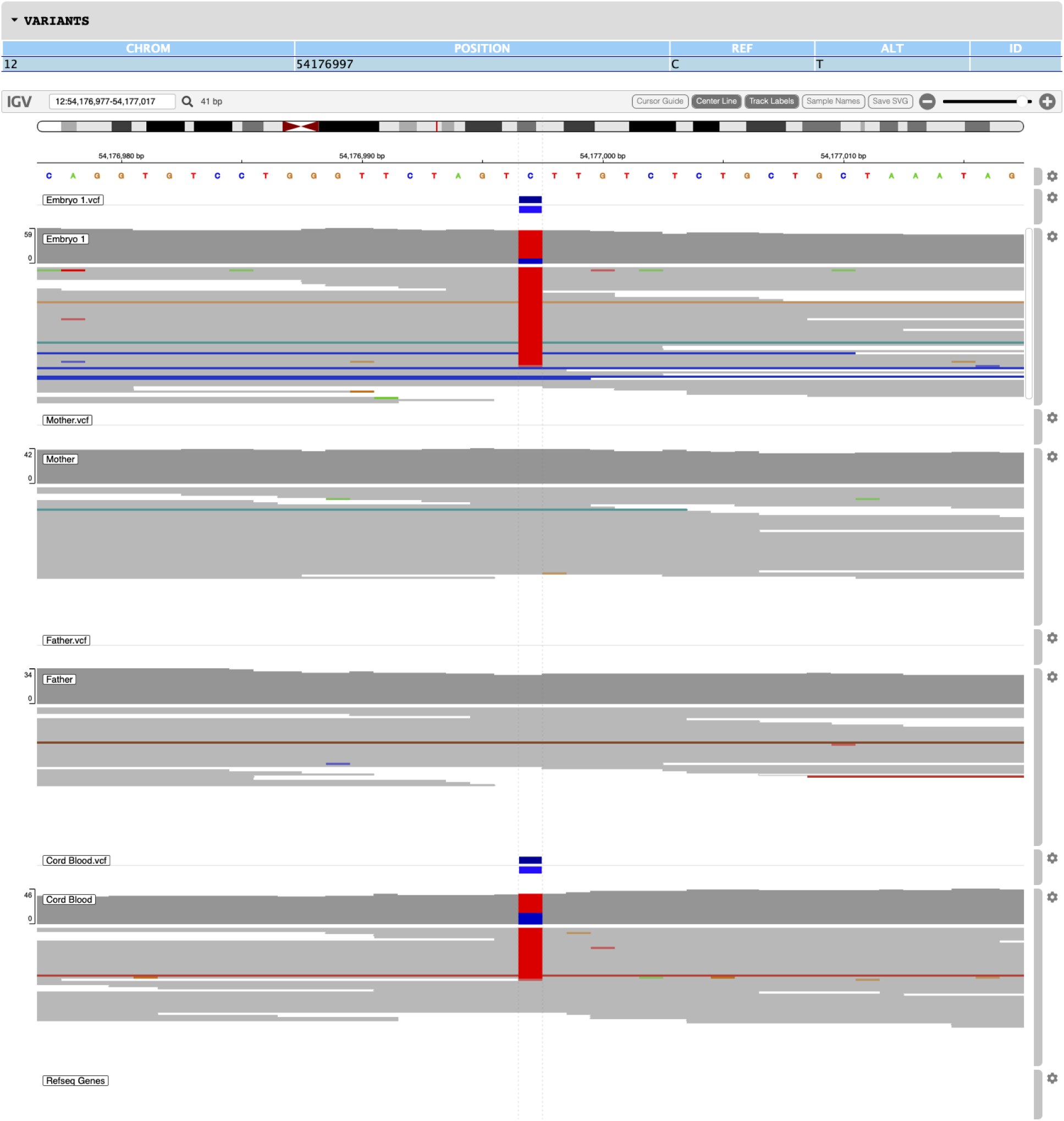
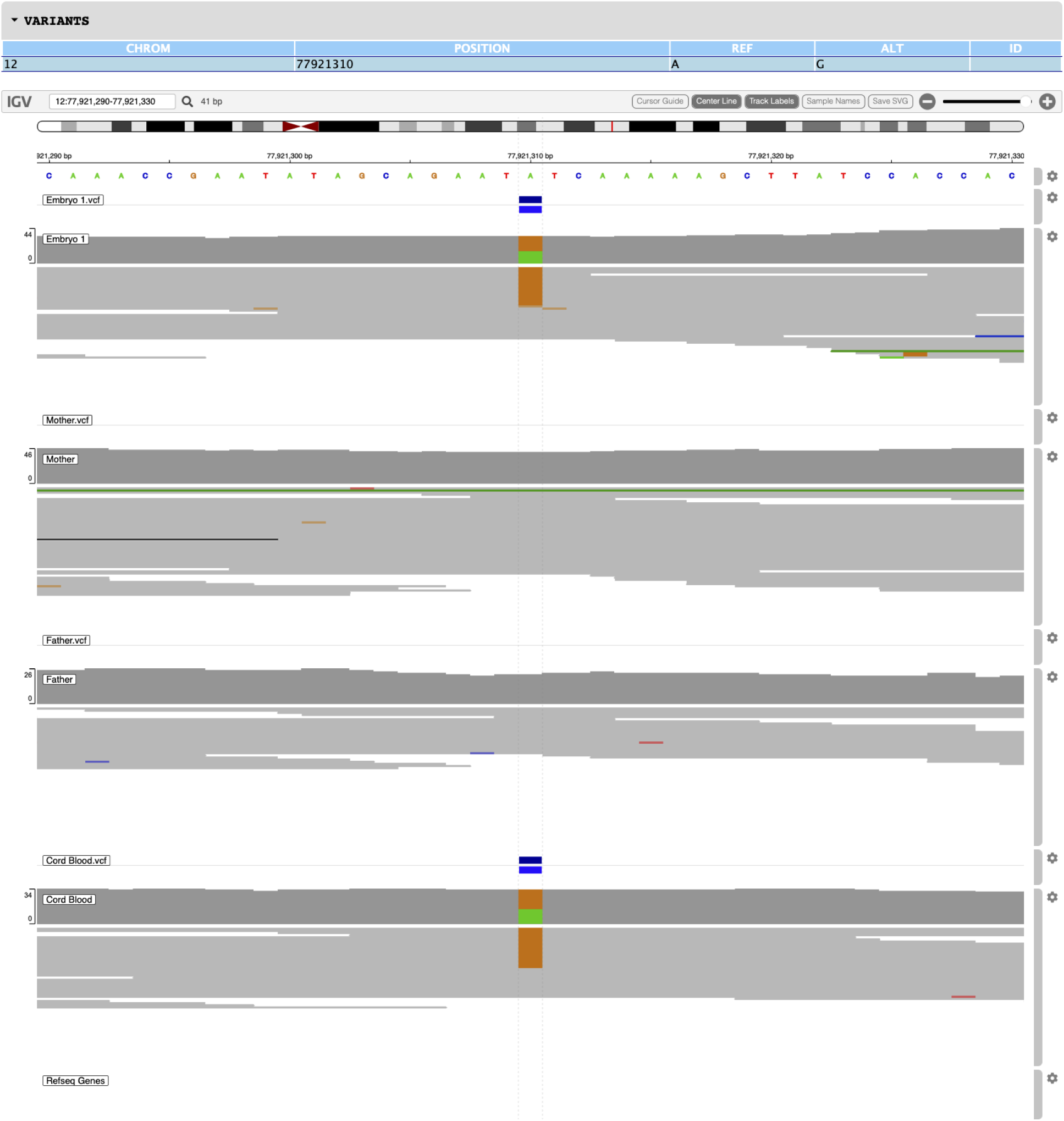
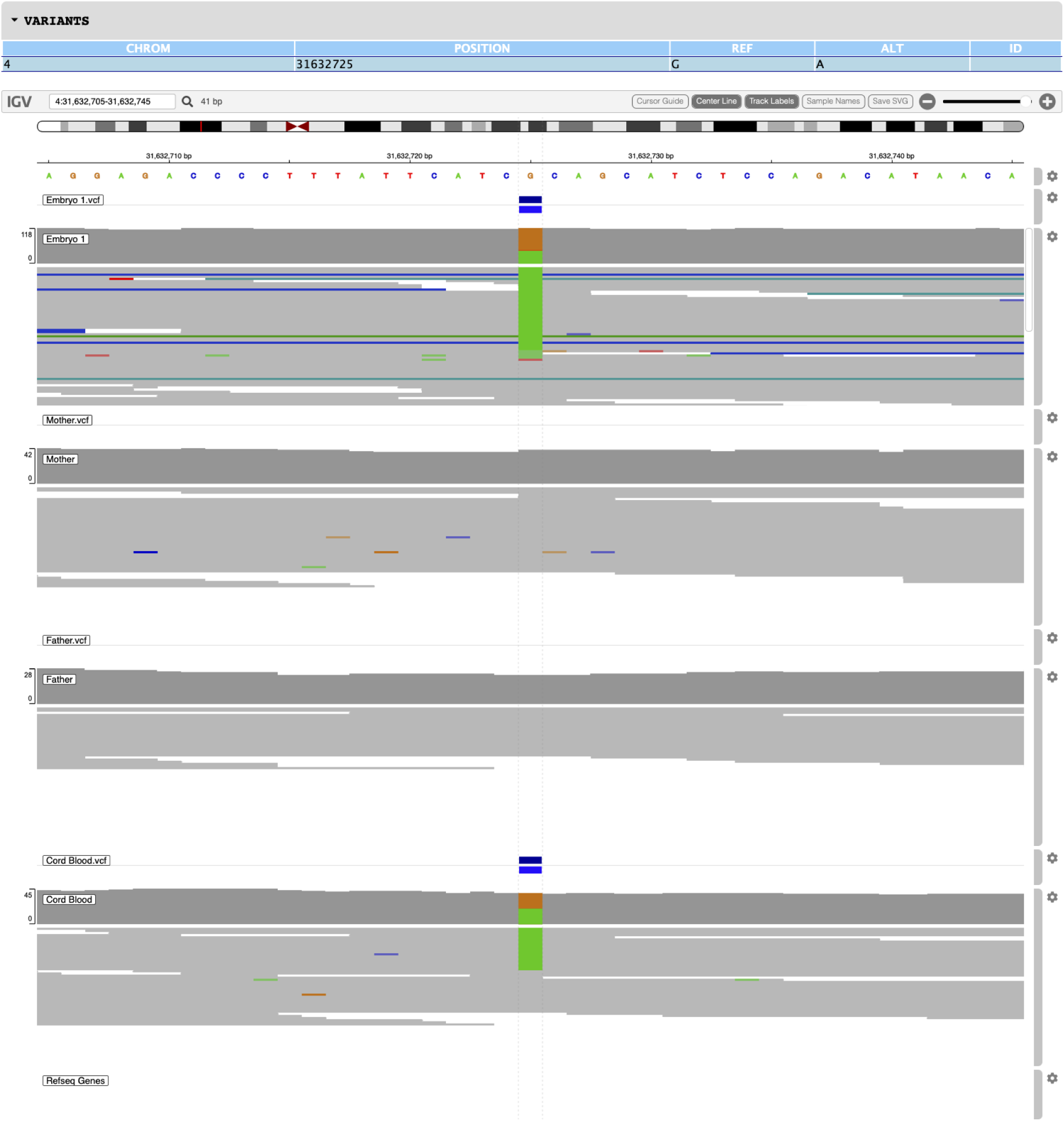
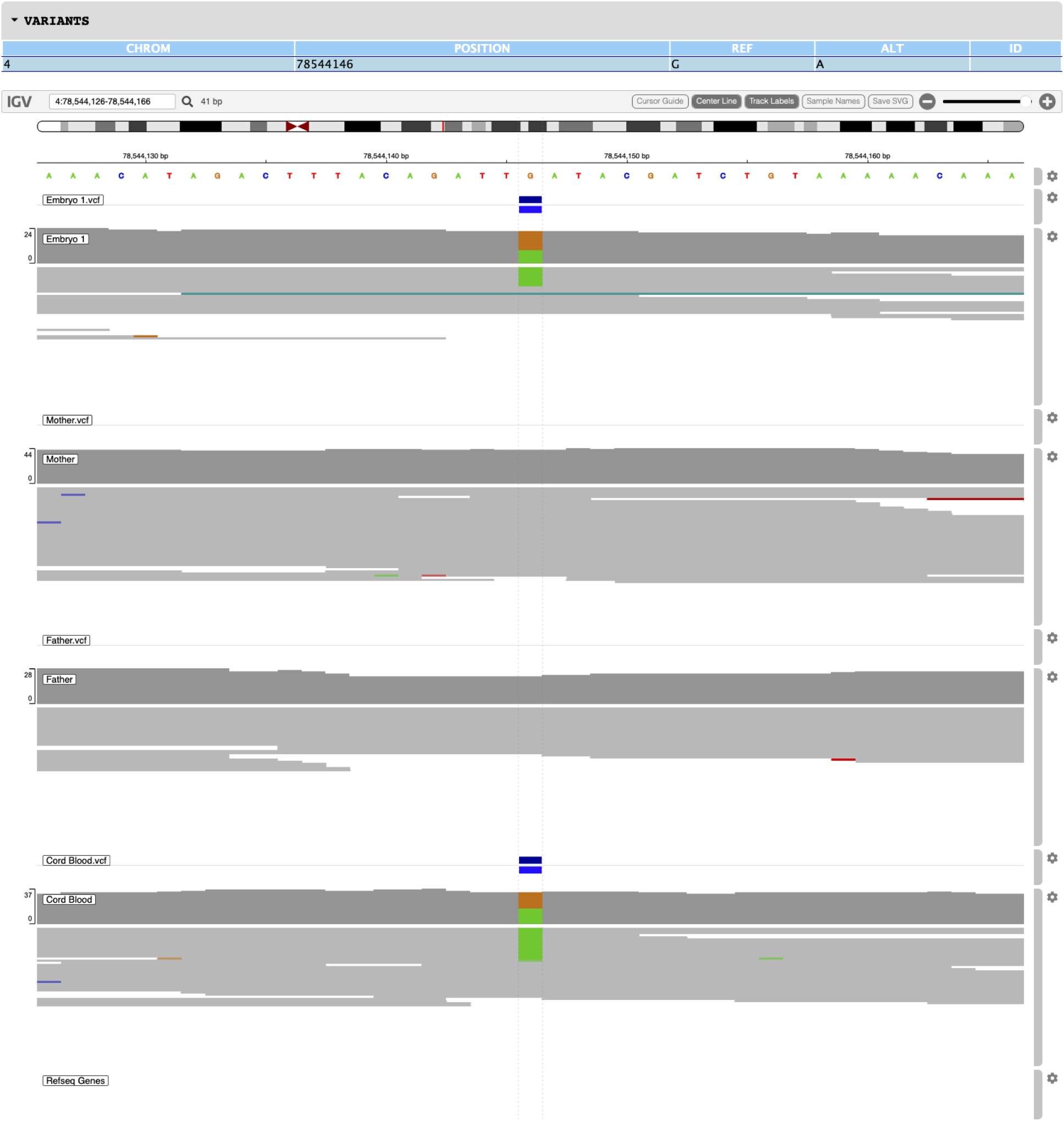
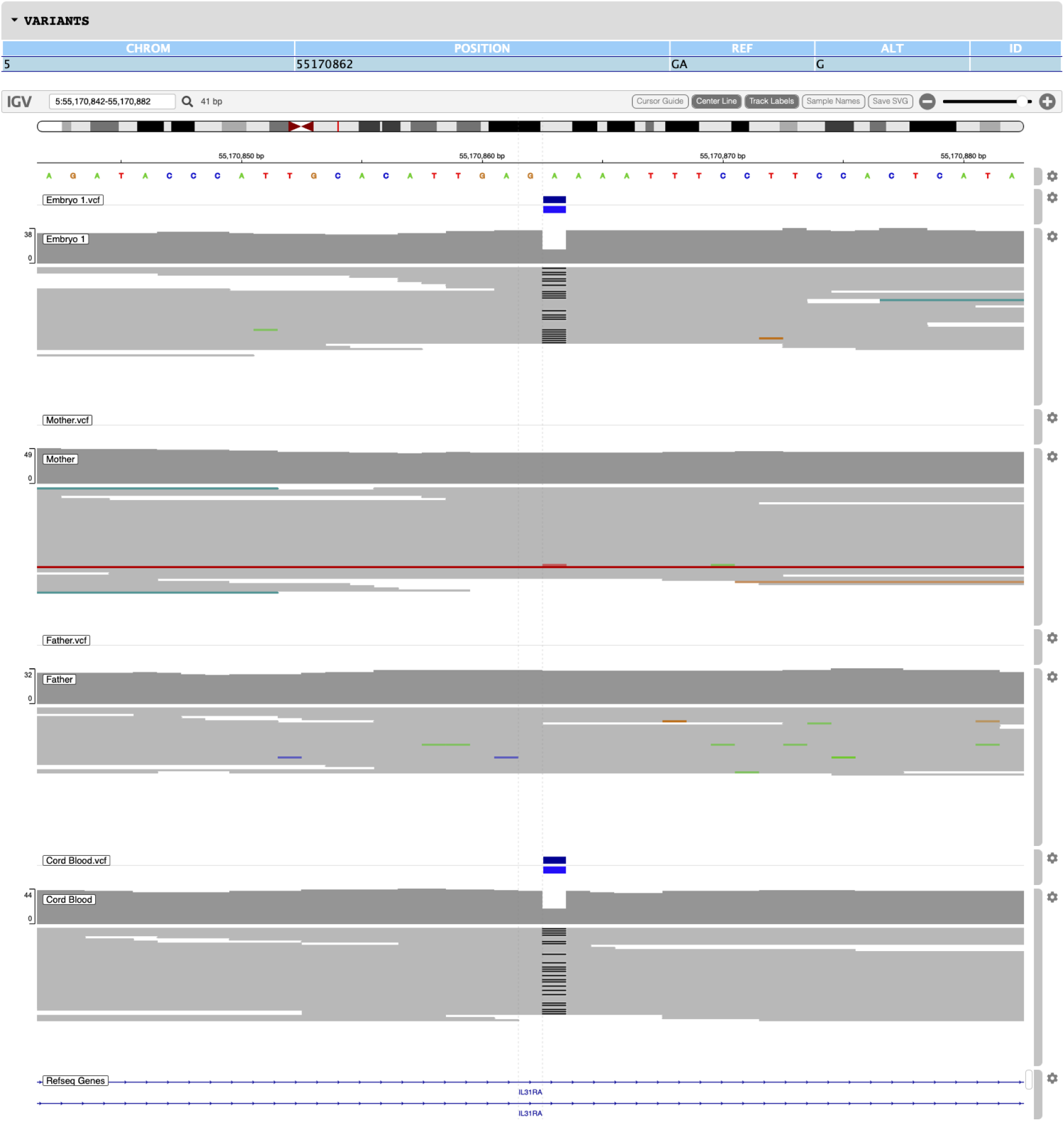

## Reference

1. Brezina, P. R., Anchan, R., & Kearns, W. G. (2016). Preimplantation genetic testing for aneuploidy: what technology should you use and what are the differences?. Journal of assisted reproduction and genetics, 33(7), 823–832. 10.1007/s10815-016-0740-2

2. Giuliano, R., Maione, A., Vallefuoco, A., Sorrentino, U., & Zuccarello, D. (2023). Preimplantation Genetic Testing for Genetic Diseases: Limits and Review of Current Literature. Genes, 14(11), 2095. 10.3390/genes14112095

3. Kraft, S. A., Duenas, D., Wilfond, B. S., & Goddard, K. A. (2019). The evolving landscape of expanded carrier screening: challenges and opportunities. Genetics in Medicine, 21(4), 790–797.

4. Lee, J., & Hong, S. E. (2019). Functional annotation of de novo variants from healthy individuals. Genomics & informatics, 17(4), e46. 10.5808/GI.2019.17.4.e46.

5. 5. Nohales, M., Coello, A., Martin, A., Insua, F., Meseguer, M., & de Los Santos, M. J. (2023). Should embryo rebiopsy be considered a regular strategy to increase the number of embryos available for transfer?. Journal of assisted reproduction and genetics, 40(8), 1905–1913. 10.1007/s10815-023-02875-z

6. Kumar, A., Im, K., Banjevic, M., Ng, P. C., Tunstall, T., Garcia, G., Galhardo, L., Sun, J., Schaedel, O. N., Levy, B., Hongo, D., Kijacic, D., Kiehl, M., Tran, N. D., Klatsky, P. C., & Rabinowitz, M. (2022). Whole-genome risk prediction of common diseases in human preimplantation embryos. Nature medicine, 28(3), 513–516. 10.1038/s41591-022-01735-0

7. Murphy, N.M., Samarasekera, T.S., Macaskill, L. et al. Genome sequencing of human *in vitro* fertilisation embryos for pathogenic variation screening. Sci Rep 10, 3795 (2020). 10.1038/s41598-020-60704-0

8. Yuntao Xia, Willy Chertman, Dhruva Chandramohan, Maria Katz, Elan Bechor, Ben Podgursky, Michael Hoxie, Qinnan Zhang, Jessica Kang, Edwina Blue, et al. First clinical validation of whole genome screening on standard trophectoderm biopsies of preimplantation embryos. bioRxiv 2022.04.14.488421; doi: 10.1101/2022.04.14.488421

9. Goldmann, J. M., Hampstead, J. E., Wong, W. S. W., Wilfert, A. B., Turner, T. N., Jonker, M. A., Bernier, R., Huynen, M. A., Eichler, E. E., Veltman, J. A., Maxwell, G. L., & Gilissen, C. (2021). Differences in the number of de novo mutations between individuals are due to small family-specific effects and stochasticity. Genome research, 31(9), 1513–1518. 10.1101/gr.271809.120

10. Acuna-Hidalgo, R., Veltman, J.A. & Hoischen, A. New insights into the generation and role of de novo mutations in health and disease. Genome Biol 17, 241 (2016). 10.1186/s13059-016-1110-1

11. 11. Deciphering Developmental Disorders Study. Prevalence and architecture of de novo mutations in developmental disorders. Nature 542, 433–438 (2017). 10.1038/nature21062

12. Chaubey, A., Shenoy, S., Mathur, A., Ma, Z., Valencia, C. A., Reddy Nallamilli, B. R., Szekeres, E., Jr, Stansberry, L., Liu, R., & Hegde, M. R. (2020). Low-Pass Genome Sequencing: Validation and Diagnostic Utility from 409 Clinical Cases of Low-Pass Genome Sequencing for the Detection of Copy Number Variants to Replace Constitutional Microarray. The Journal of molecular diagnostics : JMD, 22(6), 823–840. 10.1016/j.jmoldx.2020.03.008

13. 13. Van der Auwera, G.A., Carneiro, M.O., Hartl, C., Poplin, R., del Angel, G., Levy-Moonshine, A., Jordan, T., Shakir, K., Roazen, D., Thibault, J., Banks, E., Garimella, K.V., Altshuler, D., Gabriel, S. and DePristo, M.A. (2013), From FastQ Data to High-Confidence Variant Calls: The Genome Analysis Toolkit Best Practices Pipeline. Current Protocols in Bioinformatics, 43: 11.10.1–11.10.33. 10.1002/0471250953.bi1110s43

14. Altschul, S. F., Gish, W., Miller, W., Myers, E. W., & Lipman, D. J. (1990). Basic local alignment search tool. Journal of Molecular Biology, 215(3), 403–410. DOI: 10.1016/S0022-2836(05)80360-2

15. Koboldt, D.C. Best practices for variant calling in clinical sequencing. Genome Med 12, 91 (2020). 10.1186/s13073-020-00791-w

16. Qianqian Zhu, Qiuju Chen, Li Wang, Xuefeng Lu, Qifeng Lyu, Yun Wang, Yanping Kuang, Live birth rates in the first complete IVF cycle among 20 687 women using a freeze-all strategy, *Human Reproduction*, Volume 33, Issue 5, May 2018, Pages 924–929, 10.1093/humrep/dey044

17. Zhang, Q., Sidorenko, J., Couvy-Duchesne, B. et al. Risk prediction of late-onset Alzheimer’s disease implies an oligogenic architecture. Nat Commun 11, 4799 (2020). 10.1038/s41467-020-18534-1

18. Durtschi, J., Margraf, R.L., Coonrod, E.M. et al. VarBin, a novel method for classifying true and false positive variants in NGS data. BMC Bioinformatics 14 (Suppl 13), S2 (2013). 10.1186/1471-2105-14-S13-S2’

